# “Leveraging AI Tools to Bridge the Healthcare Gap in Rural Areas in India”

**DOI:** 10.1101/2024.07.30.24311228

**Authors:** Ajit Kerketta, Sathiyaseelan Balasundaram

**Affiliations:** Research Scholar School of Business and Management CHRIST (Deemed to be University), Bengaluru, India; Professor & Area Chair HR Specialization School of Business and Management CHRIST (Deemed to be University), Bengaluru, India

**Author notes:** **Corresponding Author:** Ajit Kerketta.

**Keywords:** Artificial Intelligence, Healthcare gap, Rural areas, remote patient monitor, healthcare outcome

## Abstract

**Introduction:** Despite considerable progress in the healthcare sector, rural regions continue to grapple with healthcare deficiencies. However, the emergence of AI technology offers promising solutions to overcome these hurdles. Hence, the study explores the potential and efficacy of introducing artificial intelligence (AI) tools to address the healthcare disparity in rural India.

**Methods:** The research employed a literature review method and gathered data from various databases such as Science Direct, PubMed, and Google Scholar. The screening process was aided by the “Rayyan” electronic software. Articles published in English between January 2020 and December 2022 were selected, followed by a thematic analysis of the findings.

**Results:** Results indicate the potential of AI in rural healthcare settings, showing AI-driven solutions addressing healthcare access gaps and contributing to their bridging. The study also highlights hurdles related to AI tool adoption in rural healthcare and proposes collaborative efforts among policymakers, healthcare providers, and technology developers to integrate AI tools effectively. This necessitates advocating for digital infrastructure investments, capacity-building initiatives, and conducive regulatory frameworks for AI implementation.

**Conclusion:** The study underscores AI’s transformative role in bridging the healthcare gap in rural India. By harnessing AI technologies, healthcare providers and policymakers can surmount barriers, empower local healthcare workers, and improve health outcomes for rural communities. The insights and recommendations contribute to the evolving knowledge base on leveraging AI for adequate healthcare delivery, guiding future initiatives in similar contexts.

## I. Introduction

Access to quality healthcare is a fundamental right.^(1–3)^ Nevertheless, basic healthcare necessities remain inaccessible to half of the global population residing in rural locales.^(4)^ India, with 68.2% of its populace residing in rural areas, faces disproportionate access to healthcare facilities. As a result, rural communities experience a healthcare gap that severely impacts their health outcomes and quality of life.^(5)^ The COVID-19 pandemic has uncovered the efficacy of the healthcare systems of India and emphasised the immediate attention for global unity in mobilising resources and leveraging innovative technologies to expedite the creation of essential healthcare solutions. However, the emergence of Artificial Intelligence (AI) presents a promising technological avenue capable of tackling these challenges and enhancing healthcare accessibility in rural regions. s‘Artificial intelligence (AI)’ is broad, longstanding, and simulates human intelligence processes by machines, particularly computer systems.^(6)^ These processes include tools such as Machine Learning, Knowledge Representation and Reasoning, Expert Systems, Robotics, and Speech Recognition. The application of AI tools has ensured resolving the deficiency of healthcare facilities and thus improved healthcare availability, accuracy of diagnosis, optimized treatment plans, improved health outcomes, and reduced costs.^(7,8)^ This research explores the potential of AI tools to bridge the healthcare gap in rural areas in India, provides an overview of the current state of rural healthcare, the potential benefits, and drawbacks of AI tools, and demonstrates findings from an empirical study assessing the effectiveness of AI tools in rural areas. The study will conclude with recommendations for future research and implementation of AI tools in rural healthcare.

### 2. Background

Rural areas are often characterised by a shortage of healthcare providers, limited healthcare facilities, and inadequate healthcare infrastructure.^(9)^ These populations have suffered from subpar healthcare facilities for decades, bearing the brunt of unequal healthcare delivery, which has led to a growing healthcare imbalance. They face barriers to healthcare access due to geographical remoteness, transportation challenges, lack of healthcare awareness, inadequate health workforce, and financial limitations.^(10)^ Nevertheless, understanding the healthcare gap in rural settings and catechising healthcare professionals on applications of Artificial Intelligence(AI) promises successful adoptions of technologies in rural healthcare systems.^(11)^ Artificial Intelligence has emerged as a promising tool with the capacity to tackle these hurdles and enhance healthcare service accessibility in rural locales.

The application of AI serves as an adjunct that benefits outcomes rather than replacement of healthcare professionals^(12)^ however, the applications of such technologies provide opportunities to ensure adequate healthcare provision.^(13)^ Implementing AI has facilitated telemedicine, allowing patients in rural areas to access healthcare remotely. Additionally, AI tools have supported diagnostic imaging and screening, enabling healthcare providers in rural areas to identify and manage health conditions more effectively. Moreover, AI has aided in optimizing healthcare logistics, including the management of medical supplies and equipment, as well as remote patient monitoring. These developments promise to reduce expenses, enhance operational efficiency, and improve patient outcomes. Numerous instances illustrate the successful integration of AI tools in rural healthcare. The study published in the Journal of Medical Internet Research confirmed that AI-based decision support tools helped improve the accuracy and efficiency of rural telemedicine consultations.^(14)^ Another study published in the Journal of Medical Systems witnessed that AI-based screening tools improved the detection of diabetic retinopathy in rural areas.^(15)^

However, within the next ten years, artificial intelligence (AI) has the potential to transform the way healthcare is delivered in rural areas by improving the accessibility, affordability, and quality of healthcare services. Therefore, the study aimed to assess the efficacy of the applications of AI tools and mitigate the healthcare disparity in rural regions in India.

## 3. Literature Review

Access to quality healthcare is a significant challenge in rural areas. The National Rural Health Association (NRHA) showed that 20% of Americans who live in rural areas have 9% of physicians.^(16)^ The discrepancy in healthcare accessibility arises from various factors, including insufficient healthcare personnel, distance from medical facilities, and inadequate funding for healthcare services in rural locales. Recent progress in artificial intelligence (AI) offered promise in narrowing the healthcare divide in rural regions. AI tools such as telemedicine, chatbots, and diagnostic algorithms hold the potential to deliver prompt and precise medical care to patients in remote areas. Telemedicine, especially, proves effective in delivering healthcare services to rural communities.^(17)^ In another study by Hollander and Carr, telemedicine provides specialty care to patients in rural areas, resulting in reduced travel time and costs for patients.^(18)^

A study found that chatbots effectively provided medical advice for minor health concerns. ^(19)^ Additionally, diagnostic algorithms based on AI can assist healthcare providers in making accurate diagnoses, particularly in areas with a shortage of healthcare professionals. ^(20)^ However, there are also concerns surrounding the use of AI in healthcare, particularly in rural areas. One concern is that AI tools may not be accessible to all patients, particularly those in low-income households or those without internet access. Another concern is that AI tools cannot replace the human touch in healthcare ^(11,12,21)^, which is important for building trust and rapport between healthcare providers and patients.

AI tools promise to bridge the healthcare disparity in rural areas; however, further research is warranted to assess their long-term impact and address accessibility and equity concerns. Healthcare providers and policymakers should explore the utilization of AI tools to enhance healthcare access in rural regions, while also acknowledging and mitigating potential challenges and concerns associated with their implementation.

### 3.1. AI Tools in Rural Healthcare

Ensuring accessible healthcare services in rural areas is a worldwide challenge, and India faces similar hurdles too. These obstacles include a shortage of healthcare professionals, lack of health awareness, inadequate infrastructure, and suboptimal healthcare outcomes. According to a report by the National Health Profile, India has one of the lowest doctor-to-population ratios in the country, with only one doctor per 5,854 people in rural areas.^19^ This shortage of healthcare workers results in long wait times and limited access to health services. Many rural health centres lack electricity, water, and sanitation facilities posing challenges for healthcare delivery.

Additionally, poor healthcare outcomes in rural areas have often been attributed to inadequate healthcare infrastructure, providers, and healthcare literacy.^(22)^ Such deficiencies have resulted in higher infant mortality, maternal mortality, malnutrition, and communicable diseases such as tuberculosis and malaria among rural communities.^(23)^

However, the Indian government has launched numerous rural health initiatives in order to bridge the healthcare gap in rural areas. These initiatives include Mobile Medical Units (MMUs), the Ayushman Bharat Digital Mission, the CoWIN App, Aarogya Setu, e-Sanjeevani, e-Hospital, e-Blood Bank, Online Registration System, Services e-Health Assistance and Teleconsultation, e-RAKTKOSH, and telemedicine, all implemented as part of the National Health Mission to enhance healthcare access in rural regions.^(24)^ However, the favour that these initiatives could offer to rural residents in accessing healthcare is still underutilized at the local level.

### 3.2. Use of AI in healthcare

Several studies have demonstrated the potential of AI-powered healthcare tools in improving healthcare outcomes in rural areas. AI tools have been shown to enhance disease detection and diagnosis, assist healthcare providers in decision-making, and improve patient outcomes. ^(25)^ For example, AI tools can assist in the early detection of diseases such as cancer and improve the accuracy of diagnoses. ^(26)^ Furthermore, AI tools help healthcare providers manage patient data, monitor patient progress, and predict patient outcomes. Some key findings from previous research are. **Clinical Decision-Making**: AI assists healthcare providers in clinical decision-making by providing real-time patient data analysis and personalised treatment recommendations. A systematic review published in the Journal of Medical Internet Research (JMIR) in 2021 found that AI-based decision support systems improved clinical decision-making in various healthcare specialties. ^(27)^ **Diagnostic Accuracy:** AI has shown great potential in improving diagnostic accuracy in various medical specialties. For instance, a study published in the Journal of the American Medical Association (JAMA) in 2020 found that an AI system outperformed human pathologists in the diagnosis of breast cancer. ^(28,29)^ **Patient Outcome Prediction**: AI can analyse large volumes of patient data to predict patient outcomes and inform treatment decisions. A study published in the Journal of Medical Informatics in 2020 found that an AI-based risk prediction model improved mortality predictions for hospitalised patients. ^(30)^ However, there are also concerns about the ethical and legal implications of using AI in healthcare, such as data privacy, algorithm bias, and accountability. Therefore, there is a need for further research to address these concerns and ensure the responsible and ethical use of AI in healthcare.

### 3.3. Challenges of AI tools in rural healthcare

AI-powered tools help reduce the workload of healthcare providers, increase efficiency in healthcare delivery, and provide patients with access to remote consultations and monitoring.^(31)^ The implementation of AI tools in rural healthcare has highlighted several potential drawbacks, including the high cost of technology, lack of technical expertise, and concerns about patient privacy and security. Rural areas often have limited resources and funding, which makes it challenging to acquire and maintain the necessary technology for AI-powered healthcare. Also, the use of AI tools requires significant technical expertise, which may be lacking in rural healthcare settings. AI-powered healthcare tools often require the collection and storage of large amounts of patient data, which are vulnerable to breaches or misuse. Patients in rural areas have limited access to legal recourse in case of a privacy or security breach. However, the application of AI tools in rural healthcare presents several potential benefits, as well as poses significant challenges that must be addressed to ensure effective and ethical use.

## 4. Methods

### 4.1. Registration and Protocol

This review article was registered with the PROSPERO International Prospective Register of Systematic Reviews (Registration Number: [CRD42023414450]). The protocol was developed according to the PRISMA (Preferred Reporting Items guidelines, ensuring a standardized and transparent review process.

### 4.2. Search strategy

The study involved a review of the existing literature on the use of AI tools in rural healthcare. The qualitative review included a systematic search of the relevant data and retrieved literature using the Google Scholar, PubMed, and Scopus (ScienceDirect) electrical databases employing appropriate keywords and search terms: “AI tools in healthcare”, “Artificial intelligence in rural healthcare”, “Machine learning”, “Deep learning”, “Remote patient monitoring” “Telehealth” “Telemedicine” “Rural health” “Rural communities” “Healthcare outcomes” “Healthcare disparities.” The review was refined by adding additional keywords and search terms: “Digital health,” “E-health”, “mHealth”, “Mobile health”, “Smart health”, and “Health technology.” The study was modified further based on its specific requirements and search syntax.

The data restriction (1st January 2020–12th December 2022) was used, and only published English-language content was extracted. The subsequent step was text availability, where an abstract was chosen, and articles with the study types of review and systematic review were added. The displayed data was elected, and the ‘Send to’ alternate was tapped, choosing “citation manager” followed by “selected all,” and a file (Pubmed-Artificial-set) nbib was created. Likewise, the author chose the ScienceDirect database from the Scopus family, refined the data retrieval by 2020–2022, and selected the review article type. All of the articles exhibited were exported to RIS format after selection. To find reliable and more relevant data, the author also used Google Scholar and conducted a manual search method. According to the author, these manual search methods—particularly hand-searching by choosing the most relevant data—led to differing study results.^(32)^

### 4.3. Inclusion Criteria

The inclusion and exclusion criteria define who can be included and excluded from the study sample. The present studies examined the use of AI tools in healthcare delivery and assessed the impact of AI tools on healthcare outcomes in rural areas. This helped identify the population in a consistent, reliable, uniform, and objective manner. ^(33)^ Studies compared the use of AI tools to other interventions or standards of care in rural areas. These included rural populations or healthcare providers as study participants. Studies published in the English language.

### 4.4. Exclusion criteria

Studies that were not centred on AI tools in healthcare delivery in rural areas. Studies that only focused on urban or suburban populations. Studies that did not report on healthcare outcomes or the impact of AI tools on healthcare outcomes. Studies that only reported on technical aspects of AI tools without examining their impact on healthcare outcomes. Studies published in languages other than English. In addition, the criteria should be clearly defined and applied consistently throughout the study to ensure that the study is rigorous and reliable.

### 4.5. Study selection

Data screening is a crucial step after the data search. However, retrieved articles were screened by two reviewers using an AI-powered application called “Rayyan”, which allowed authors to work more accurately and quickly. The initial abstract and title screening process was sped up using Rayyan software, which combined semi-automation with excellent usability.^(34)^ Of the searches, 1008 records were migrated to Rayyan. The research title was designed before the data was uploaded. Data auto duplication was used after uploading and discovered two exact matches, which were resolved in the process. The screening tool included four inclusion decision categories: “Undecided,” “Maybe,” “Included,” and “Excluded.” The complete screening process of the study was done in three phases: The author glanced through the title and abstract for the preliminary phase. The study’s title contained the terms “Artificial Intelligence,” “Rural/Healthcare,” “Machine learning,” and “Application of AI,” which were all included in the inclusion decision with the automatically generated justification “Background article” and given the label “AI.” The ambiguous data for which the author assumed a chance of finding relevant information was labeled as “P” and placed in the “Maybe” category. Additionally, the information deemed inappropriate and irrelevant to the subject was categorically eliminated and labeled “Irrelevant.” In the second phase, two titles had to be created: one for the data classified as “Maybe,” with the title “Maybe full-text review,” and another for the data classified as “Included,” with the title “Included full-text review.” The data that are most pertinent to the topic were categorised for the final full-text study for the third and last phases of the screening process.

### 4.6. Data extraction

The study was selected on the basis that it examined the use of AI tools in healthcare delivery in a study that evaluated the impact of AI tools on health outcomes in rural areas. This involved data extraction from the selected studies encompassing artificial intelligence and the healthcare sector, published between 2019 and 2022. One of the key elements that qualified the study for additional research was the study’s conclusion. This information was recorded in a data extraction form (WinRAR ZIP archive).

### 4.7. Quality Assessment

The level of evidence of the chosen studies was evaluated to assess the overall quality of the evidence and the risk of bias. This was done using a qualitative study approach; hence the Joann Briggs Institute Critical Appraisal Checklist ^(35)^ was employed as the relevant tool.

### 4.8. Data charting process

The study was conceptualized on April 5, 2023, and data was extracted from electronic databases, including Science Direct, PubMed, and Google Scholar, between April 15 and May 15, 2023. A total of 1009 data points were archived in the ‘Rayyan’ software for screening on May 18, 2023. After screening all the data on June 3, 2023, 28 pieces of literature were marked as ‘Maybe’ and archived in Rayyan. On June 4, 2023, three additional data points were added, and 24 ‘final full-text’ pieces were marked and archived in Rayyan on July 7, 2023. Upon screening these 24 pieces, 14 were deemed relevant to the study’s objectives, and a conclusion was drawn based on these 14 datasets. Drafting and writing of the research began in August 2023, and after numerous revisions, the study was completed on June 26, 2024, as indicated in the PROSPERO anticipated completion date.

### 4.9. Data synthesis

The study employed a qualitative approach to synthesize data. The author thoroughly reviewed the data iteratively to discern patterns, trends, and recurring themes. A systematic coding method was utilized to segment the data into meaningful units, resulting in the generation of initial codes. Subsequently, these codes were organized into relevant themes through clustering, forming primary themes. The researcher meticulously examined the interplay between codes and themes, refining and adjusting them as necessary to ensure logical coherence and consistency. Themes were iteratively developed through an inductive process of data-driven analysis, involving continuous comparison and contrast of different data segments to identify commonalities and variations. This iterative process culminated in the creation of final themes, which were presented in a cohesive and descriptive manner.

## 5. Results

### 5.1. Synthesis of results

The study employed a qualitative approach to synthesize data. The author thoroughly reviewed the data iteratively to discern patterns, trends, and recurring themes. A systematic coding method was utilized to segment the data into meaningful units, resulting in the generation of initial codes. Subsequently, these codes were organized into relevant themes through clustering, forming primary themes. The researcher meticulously examined the interplay between codes and themes, refining and adjusting them as necessary to ensure logical coherence and consistency. Themes were iteratively developed through an inductive process of data-driven analysis, involving continuous comparison and contrast of different data segments to identify commonalities and variations. This iterative process culminated in the creation of final themes, which were presented in a cohesive and descriptive manner.

The above displayed in Figure 1, (PRISMA 2020 diagram) indicates that the data identified from the electronics database n=1009 had n=4 records removed before screening for duplication and n=1 records marked as ineligible by automation tools, resulting in a total of n=1008 records screened. The full text of n=24 records was assessed for eligibility but excluded by the Rayyan Intelligent Systematic Review software for the following reasons:

- Reason 1: 3 records had the wrong study outcomes.
- Reason 2: 2 records had the wrong study design.
- Reason 3: 1 record was the wrong publication.

**Figure 1:**
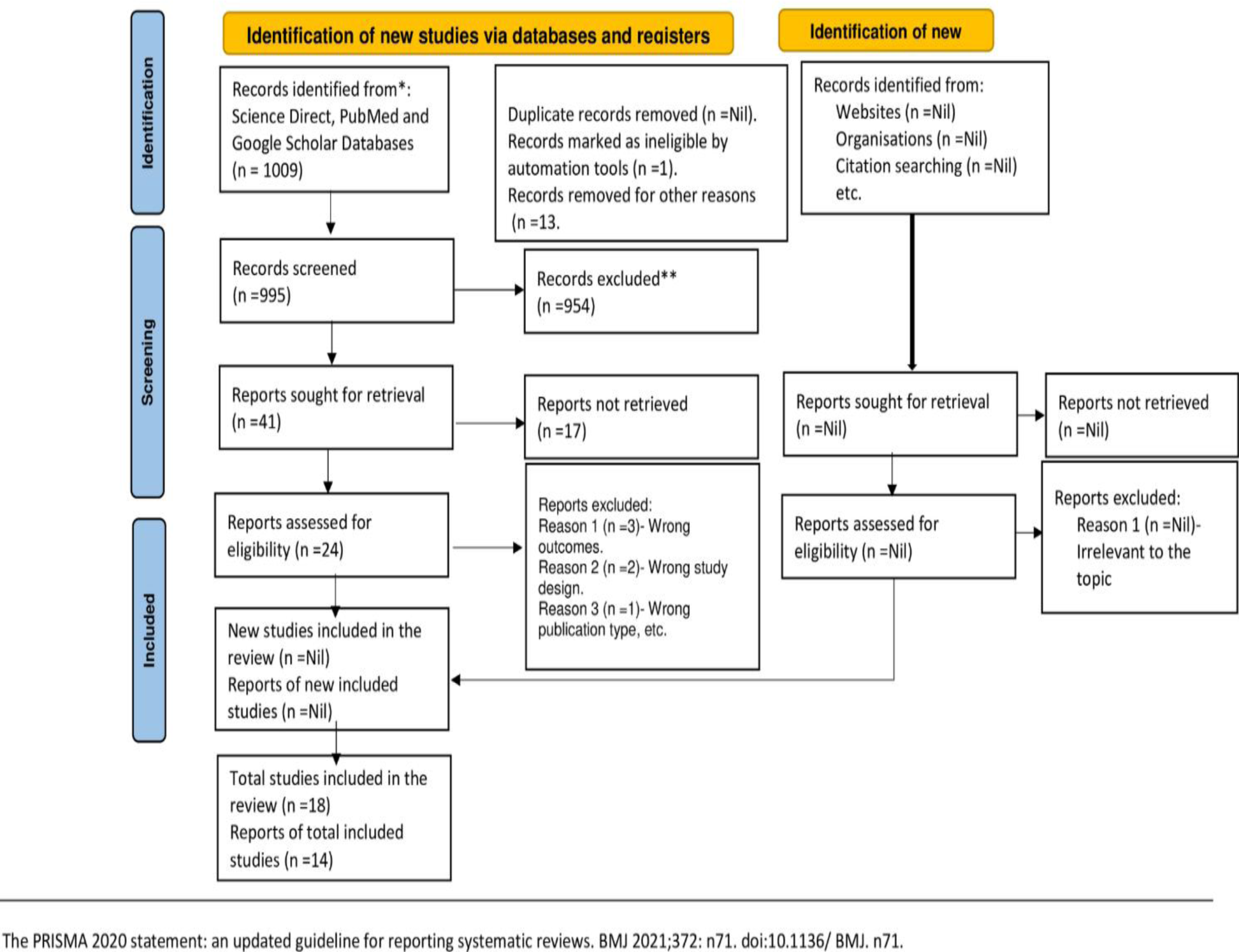
PRISMA flow diagram.

Further, categorising the objectives of the study shown in Figure 1 illustrate the notable emphasis (20.34%) on employing AI technologies to improve healthcare provision in rural regions. Addressing the rural healthcare gap emerges as the primary focus of the study, representing nearly half (44.07%) of the findings. Additionally, the study dedicates 25.42% of its outcomes to investigating the challenges associated with integrating AI tools into rural healthcare settings, accounting for over a quarter of the study’s results.

**Table 1**: Demonstrates *“Integration of AI in Healthcare”* displayed Column 1 (hereafter referred to as C1) illustrates various types of AI tools. Column 2 (C2) showcases the feasibility of AI in low-resource settings, signified by a percentage of 25%. Column 3 (C3) highlights the effectiveness of AI in rural India, displayed with a percentage of 35%. Column 4 (C4) discusses the challenges encountered in the application of AI in rural India, identified as significant impediments. The statistics presented in Table 1 illustrate various categories of AI tools aimed at enhancing rural healthcare (C1). However, their feasibility for implementation in these areas is shown to be 25% (C2). Despite this, their effectiveness in addressing the rural healthcare gap is indicated at 35% (C3). On the other hand, the challenges associated with adopting AI tools in rural regions are higher, at 40% (C4).

**Table 1:**
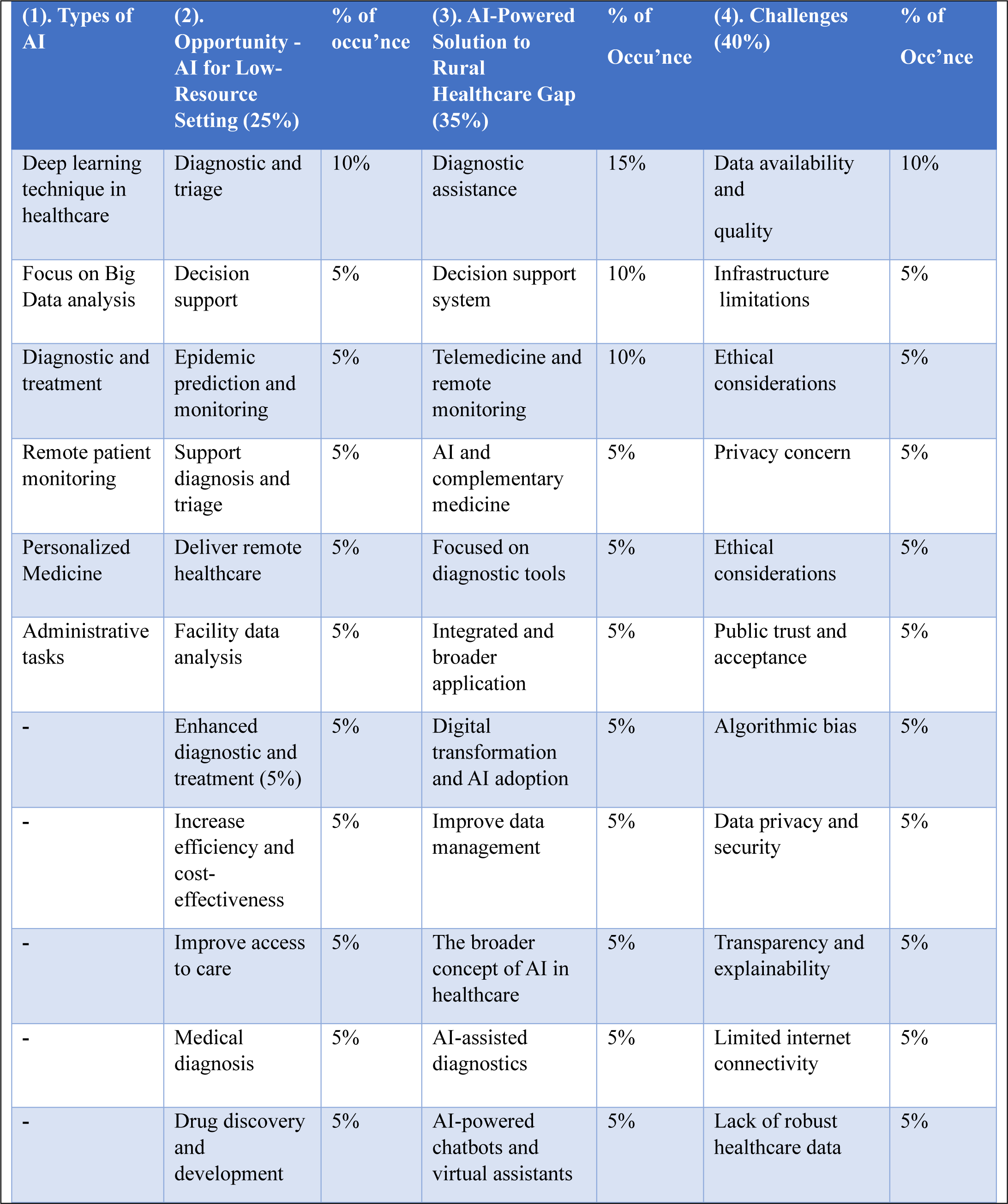
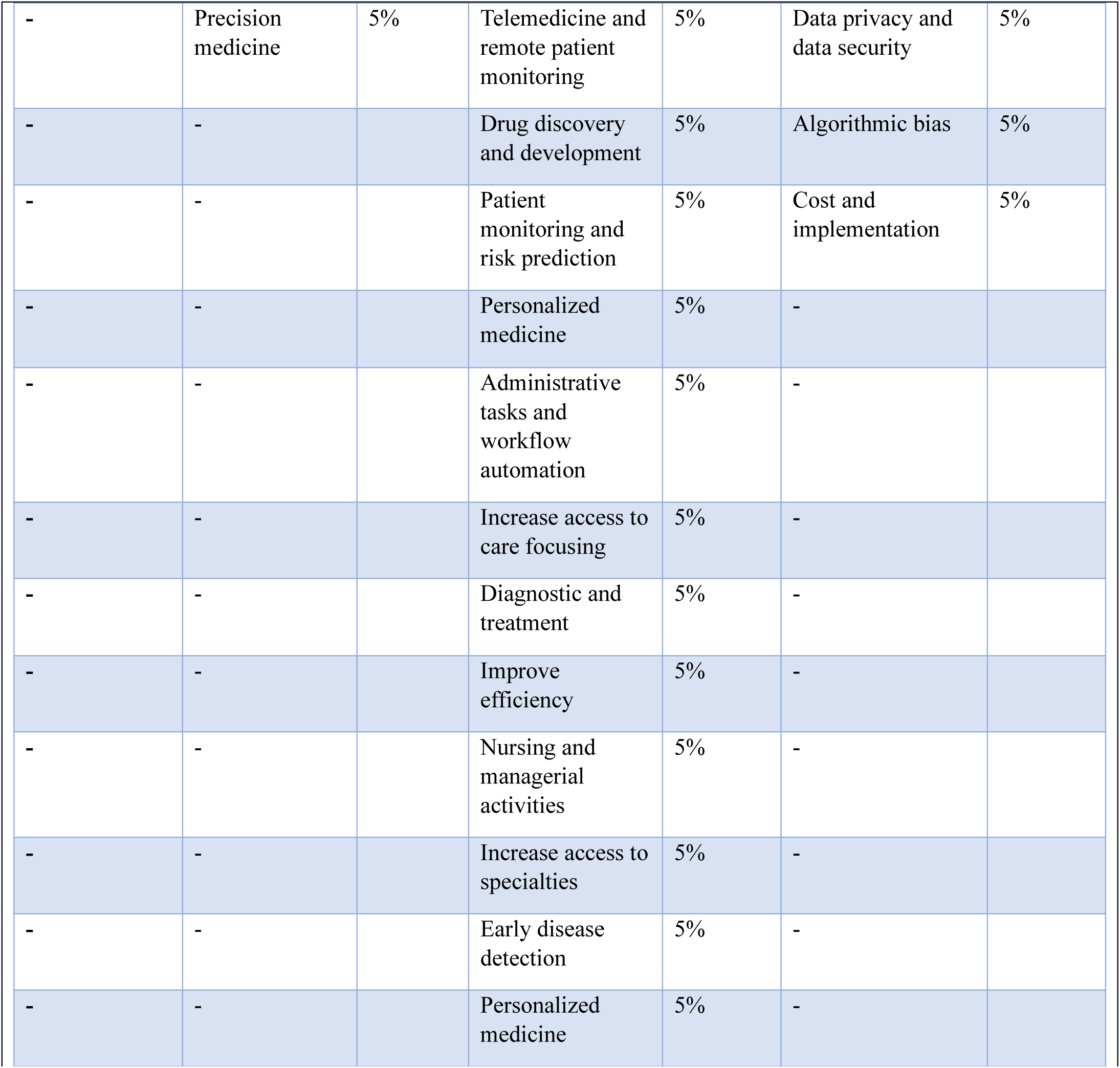
“Integration of AI in Healthcare: Opportunities, Solutions, and Challenges in Low-Resource and Rural Settings”.

| (1). Types of AI | (2). Opportunity - AI for Low-Resource Setting (25%) | % of occurrence | (3). AI-Powered Solution to Rural Healthcare Gap (35%) | % of Occurrence | (4). Challenges (40%) | % of Occurrence |
| --- | --- | --- | --- | --- | --- | --- |
| Deep learning technique in healthcare | Diagnostic and triage | 10% | Diagnostic assistance | 15% | Data availability and quality | 10% |
| Focus on Big Data analysis | Decision support | 5% | Decision support system | 10% | Infrastructure limitations | 5% |
| Diagnostic and treatment | Epidemic prediction and monitoring | 5% | Telemedicine and remote monitoring | 10% | Ethical considerations | 5% |
| Remote patient monitoring | Support diagnosis and triage | 5% | AI and complementary medicine | 5% | Privacy concern | 5% |
| Personalized Medicine | Deliver remote healthcare | 5% | Focused on diagnostic tools | 5% | Ethical considerations | 5% |
| Administrative tasks | Facility data analysis | 5% | Integrated and broader application | 5% | Public trust and acceptance | 5% |
| - | Enhanced diagnostic and treatment (5%) | 5% | Digital transformation and AI adoption | 5% | Algorithmic bias | 5% |
| - | Increase efficiency and cost-effectiveness | 5% | Improve data management | 5% | Data privacy and security | 5% |
| - | Improve access to care | 5% | The broader concept of AI in healthcare | 5% | Transparency and explainability | 5% |
| - | Medical diagnosis | 5% | AI-assisted diagnostics | 5% | Limited internet connectivity | 5% |
| - | Drug discovery and development | 5% | AI-powered chatbots and virtual assistants | 5% | Lack of robust healthcare data | 5% |
| - | Precision medicine | 5% | Telemedicine and remote patient monitoring | 5% | Data privacy and data security | 5% |
| - | - |  | Drug discovery and development | 5% | Algorithmic bias | 5% |
| - | - |  | Patient monitoring and risk prediction | 5% | Cost and implementation | 5% |
| - | - |  | Personalized medicine | 5% | - |  |
| - | - |  | Administrative tasks and workflow automation | 5% | - |  |
| - | - |  | Increase access to care focusing | 5% | - |  |
| - | - |  | Diagnostic and treatment | 5% | - |  |
| - | - |  | Improve efficiency | 5% | - |  |
| - | - |  | Nursing and managerial activities | 5% | - |  |
| - | - |  | Increase access to specialties | 5% | - |  |
| - | - |  | Early disease detection | 5% | - |  |
| - | - |  | Personalized medicine | 5% | - |  |

Figure No. 2. “Percentage Distribution: AI’s Role in Rural Healthcare - Solutions and Challenges”

**Figure 2.**
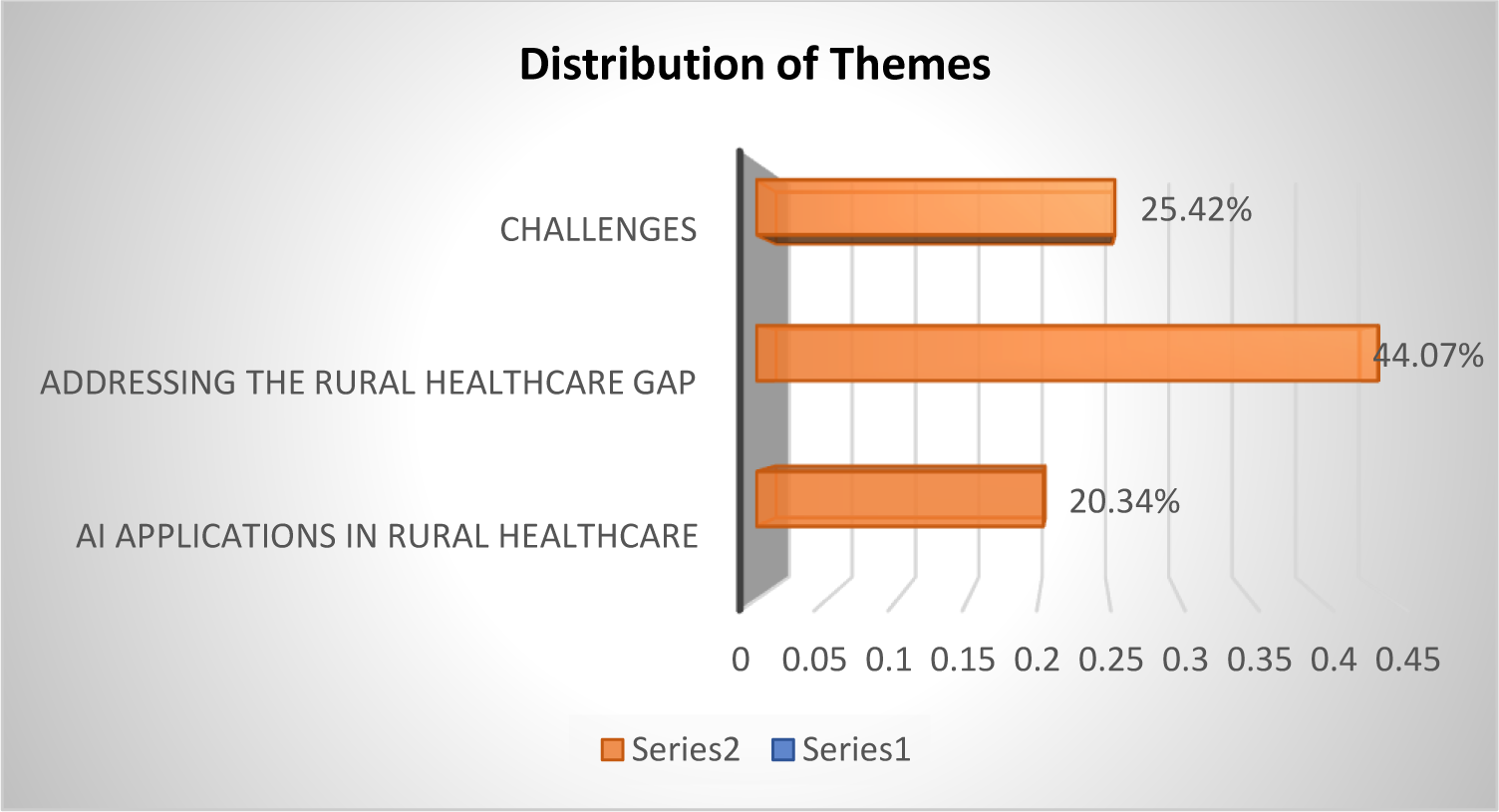
Percentage of Themes Distributed in Rural Healthcare.

### 5. 2. AI applications in rural healthcare

A significant portion, 20.34%, of the study’s outcomes emphasised employing AI tools to improve rural healthcare facilities. The study demonstrated the use of AI technologies has enhanced diagnostic capabilities, offered decision support, predicted and monitored epidemics, delivered remote healthcare services, improved efficiency and cost-effectiveness, and advanced precision medicine, among other opportunities. Within this overarching theme, several subthemes have surfaced:

#### Advanced Diagnostics and Treatment

The numeric values represent a merging of various codes contributing to this overarching theme. However, the advanced diagnostics and treatment involved leveraging deep learning and AI tools for diverse diagnostic tasks, including image analysis, providing support for treatment planning, and potentially aiding in surgical procedures.(36)

#### Improved Patient Care

The theme highlighted the impact of AI in remote patient monitoring, facilitating early issue detection, personalized medicine strategies, and potentially bolstering mental health support.(37)

#### Operational Efficiency

The operational efficiency delves into the utilization of AI for automating administrative tasks, optimizing workflows, and enhancing the overall efficiency of healthcare systems.(38)

#### Drug Discovery and Development

AI to accelerate the processes of discovering and developing new drugs.(39,40)

#### Broader Applications

This included a range of potential AI applications in healthcare, such as decision support systems, epidemic prediction, telemedicine, integrating AI into broader healthcare systems, and supporting complementary medicine. These findings provide valuable insights into the transformative impact of AI on healthcare delivery, offering valuable implications for policymakers, healthcare providers, technology developers, and stakeholders invested in advancing healthcare equity and improving patient outcomes in rural communities.

### 5. 3. Addressing the rural healthcare gap

The largest portion, accounting for approximately 44.07%, of the study focused on exploring AI-powered solutions specifically aimed at bridging the healthcare gap in rural areas. The subsequent components explored in-depth show how AI tools facilitated the bridging of these gaps in rural healthcare:

#### AI-powered Solution

The integration of AI-powered solutions has revolutionized healthcare delivery in these underserved regions, contributing significantly to bridging the healthcare gap. This illustration highlighted Telemedicine, remote patient monitoring, administrative tasks, workflow automation, increased access to care focusing, increased access to specialties, and Early disease detection as key ways in which AI technologies have transformed rural healthcare and improved health outcomes. ^(41)^

#### Improved Rural Healthcare Delivery

This theme highlighted how AI directly improved healthcare delivery in rural areas by aiding diagnostics, treatment, and streamlining operational efficiency. ^(42)^

## 6. Discussion

The study aimed to explore the impact of AI implementations in rural healthcare in India. However, it discovers a positive outcome in a different geographical setting, demonstrating how AI applications enhanced the accessibility of quality healthcare in rural areas. The study published in BMJ Global Health emphasized that the effectiveness of AI solutions in under-resourced contexts hinges on understanding local dynamics, defining clear usability criteria, and accessing sufficient training through field trials. ^(43)^ Also, the study featured in Frontiers highlighted that when implementing AI in low-resource settings, it is best to use iterative, field-based processes, that are built into existing systems and institutions rather than starting from scratch or hoping to replace existing systems, however, broken-institutions that cannot fix itself is likely to be able to support and use a complex technology properly.^(6)^ However, AI adoption facilitated the identification and evaluation of diverse AI applications customized for rural healthcare settings in India. Likewise, the present study highlighted precise AI applications, such as advanced diagnostics, remote patient monitoring, and operational efficiency tools, that have the potential to enhance accessibility and quality healthcare delivery in rural areas. Again, another study published in Health Equity titled, “The application of AI technology in rural areas of developing countries” explored that the implementation of AI provided guidance and recommendations for nurse and paramedical staff. The study also showed that the overall rate of consistency between the early detection and prevention system (EDPS) and physicians was 94%. The study also revealed that patient responses were positive following the implementation of AI technology. As a result, village health nurses were interested in using electronic devices such as mobile, laptops, iPads, and other wearable devices which enabled healthcare workers to improve the quality of healthcare delivery in rural areas. ^(44)^

The applications of AI in rural India align with the objectives of identifying and evaluating AI solutions for rural healthcare, providing valuable insights into how technology can address healthcare disparities. One of the healthcare centres in Bangalore, India, has employed cloud-computing medical services and extended its reach to remote areas in northern Indian states is one such example. Through this initiative, nurses and paramedical staff stationed in rural areas received comprehensive training to effectively manage various medical assistance. Meanwhile, skilled and qualified physicians remotely monitor the operations from Bangalore. A chapter titled “Remote Patient Monitoring using IoT, cloud-computing, and AI” from the book “Hybrid Artificial Intelligence and IoT in Healthcare” illustrated that AI has improved healthcare accessibility and quality in rural areas through remote patient monitoring using IoT, cloud computing, and other AI tools. ^(45)^ Thus, the outcomes of the study addressed the research question, “How AI be leveraged to improve healthcare delivery in rural areas? These applications align with the objectives of the study.

The study also identified potential hurdles and constraints associated with deploying AI tools in rural healthcare settings in India. When it comes to rural healthcare, incorporating artificial intelligence (AI) poses the following distinctive challenges:

### Data Issues

This theme acknowledged the challenges of limited data availability ^(46)^, quality, and robustness, especially in rural settings, which can hinder AI development and implementation in healthcare.

### Ethical Considerations

The study emphasized the importance of addressing ethical issues ^(47)^ like privacy concerns, algorithmic bias, transparency, and public trust when deploying AI in healthcare.

### Infrastructure Limitations

This study demonstrated the challenges of limited internet connectivity in rural areas, which can impede the adoption of AI-based healthcare solutions. ^(48–50)^

### Cost and Implementation

This theme acknowledged the need to consider the cost-effectiveness and practicalities of implementing AI solutions in healthcare settings, particularly in resource-constrained environments. ^(51)^

## 7. Theoretical and Practical Implications

The study contributes to the theoretical understanding of how AI tools can be leveraged to address healthcare disparities in rural areas. It expands the existing literature by exploring the potential of AI applications in the Indian context, adding to the body of knowledge on AI in healthcare. The study provides theoretical insights into the challenges and opportunities associated with implementing AI tools in rural healthcare settings. By examining factors such as infrastructure limitations, data privacy concerns, and cultural considerations, it offers valuable insights for future research on AI implementation.

The findings of the study inform the development of theoretical frameworks for evaluating the effectiveness and feasibility of AI interventions in rural healthcare. This guides future research and policy development in the field of AI and healthcare access.

## 8. Practical Implications

The study offers practical recommendations for policymakers, healthcare providers, and technology developers to harness the full potential of AI in bridging the healthcare gap in rural India. These recommendations can inform policy decisions aimed at improving healthcare access and outcomes in rural areas. The study highlights the importance of capacity-building initiatives to empower local healthcare workers and facilitate the adoption of AI tools in rural healthcare settings. Policymakers and healthcare organizations can use this information to design training programs and skill-building initiatives for healthcare professionals. The findings of the study can inform the development of AI-based healthcare solutions tailored to the needs and challenges of rural communities in India. Technology developers use this information to design and deploy AI tools that address the deficiency of healthcare gaps in rural areas and improve access to quality care.

## 8. Limitations

The study may be limited by the availability and quality of healthcare data from rural areas, which can be inconsistent, incomplete, or inaccurate. The methods used in the study may not adhere to universally accepted standards, however, followed the guidelines for the PRISMA 2020 checklist.

## 9. Conclusion and Recommendations

The study highlighted the significant potential of AI applications in improving healthcare delivery and addressing disparities in rural regions. Through an exploration of various AI tools tailored for rural healthcare settings, including advanced diagnostics and remote patient monitoring, the study underscored the transformative impact AI brought on healthcare outcomes. Despite the positive implications, significant challenges such as restricted data accessibility, ethical considerations, and infrastructure limitations posed considerable concerns and must be addressed to ensure successful implementation. Moreover, the researcher from rural India emphasized that operational challenges arise due to factors such as geographic distribution, socioeconomic constraints, and illiteracy among significant segments of the population.

However, the study also posited the following recommendations, which might significantly improve the delivery of healthcare in rural areas: Improve Data Accessibility, Set Ethical Standards, Develop Infrastructure, Offer Education and Training, Run Pilot Projects, and Promote Cooperation.

## Annexure 1

- The author has not received any sort of funding
- The author has no competing interest
- Author’s contributions:

### Author^1^

Conceived and designed the analysis

Collected the data

Analysed the data

Perform the analysis

Drafted the paper

### Author^2^

Provided technical support

Reviewed the study

## Annexure 2

### Supporting documents

1. Full text included for analysis
2. PRISMA 2020 flow diagram
3. Initial coding

## Data Availability

All data produced in the present study are available upon reasonable request to the authors

